# Assessing Health Facility Capacity for Neglected Tropical Diseases in a Refugee Settlement in Uganda

**DOI:** 10.1101/2025.10.16.25338139

**Authors:** Elvis Tamale, Paddy Derrick Malinga, Mable Ssebaduka Nabweteme, Celine Ahurira, Conrad Makai, Kenneth Ssembuze, Patricia Ajeru, Patience Atuhaire

**Affiliations:** St Mary’s Hospital Lacor, Gulu city, Uganda; Mbarara University of Science and Technology, Mbarara, Uganda; Medical Teams International, Portland, USA; Kampala International University, Kampala, Uganda; Makerere University Johns Hopkins University Research Collaboration, Kampala, Uganda

**Keywords:** Neglected Tropical Diseases, Refugee Settlement, Uganda, Primary Healthcare, Health Workers, Knowledge, Capacity, Schistosomiasis, Soil-Transmitted Helminths

## Abstract

**Background:** Neglected Tropical Diseases (NTDs) pose a substantial global public health challenge, particularly in vulnerable refugee populations. In Uganda, millions are at risk, yet primary healthcare (PHC) facility capacity within refugee settlements to effectively manage these diseases remains underexplored. This study assessed infrastructure, diagnostic capabilities, and resource availability for NTD management in health facilities across Nakivale Refugee Settlement, Uganda.

**Methods:** A quantitative assessment was conducted across 8 health facilities (6 Health Centre III, 2 Health Centre IV) in Nakivale Refugee Settlement. Data collection included facility type, bed capacity, dedicated laboratory services and personnel, availability of diagnostic tests and guiding tools, status of reporting systems, and essential medicines/laboratory supplies for soil-transmitted helminths (STH) and schistosomiasis. Descriptive statistics summarized findings.

**Results:** The 8 assessed facilities (6 HCIII, 2 HCIV) averaged 23 and 50 beds, respectively. All facilities reported dedicated laboratories with technicians and microscopes. Diagnostic tests (microscopes, dyes, assay strips) and guiding tools (clinical guidelines, laboratory protocols, bench aids) were consistently available, though specific test types varied. Five out of eight (62.5%) facilities had Health Management Information System reporting forms. Essential medicines, Albendazole and Mebendazole, were available in most facilities (7 had both, 1 had Albendazole only). Common laboratory supplies (e.g., centrifuge tubes, syringes) were universal, but critical items like cellophane and Kato Katz templates were absent in all facilities. Most facilities were over 10 km from the town council.

**Conclusion:** Health facilities in Nakivale Refugee Settlement possess foundational infrastructure for NTD management, including laboratories, microscopes, and essential medicines. However, significant gaps exist in specific diagnostic supplies (e.g., Kato Katz templates, cellophane) and reporting system completeness. Their predominantly remote location also highlights potential access challenges. Addressing these gaps through targeted provision of laboratory consumables, strengthening reporting, and considering geographical accessibility is crucial for enhancing PHC capacity to manage NTDs in this refugee setting.

## Introduction

Neglected tropical diseases (NTDs) are a group of 13 infections caused by parasitic worms, protozoa, or bacteria currently affecting over 1 billion people, with another 1 billion at risk of infection [1–8, 25–31, 33]. They strike the world’s poorest people with adverse effects on the health, well-being, and socioeconomic facets of one’s life [2, 9–11, 34]. Africa bears a substantial portion of this global burden, accounting for over 600 million cases, particularly affecting poor rural communities [1–2, 12, 36]. Uganda is endemic for all five NTDs, with more than 40 million people at risk for one or more NTDs targeted by the United States Agency for International Development (USAID) Act to End NTDs [13, 37].

Despite their significant burden, NTDs are largely preventable and amenable to elimination. In 2020, the World Health Organization (WHO) outlined a roadmap for eradicating and eliminating 20 NTDs by 2030 [14, 38]. The WHO, alongside policymakers and academics, advocates for integrating NTD management into existing public health programs within endemic countries [39, 40]. Such integration is posited to contribute significantly to achieving several sustainable development goals (SDGs) and vice versa [14–16, 32, 35]. Despite the burden of NTDs in Sub-Saharan Africa, NTDs have not been well integrated into the PHC of most countries. Previous studies in Nigeria, Tanzania, and Burundi have consistently reported low capacity and readiness among health workers to diagnose and manage NTDs [1, 17–18, 41, 42]. The Ministry of Health in Uganda has identified several hindrances to eliminating NTDs by 2030, including a lack of knowledge, skills, and equipment for diagnosis and management in health facilities and deficiencies in reporting systems [23].

Nakivale Refugee Settlement, the eighth-largest settlement globally, hosts approximately 145,613 refugees from ten different nationalities [19, 43]. Within this settlement, NTDs, particularly schistosomiasis and soil-transmitted helminths (STHs), are prevalent, with reported rates of 26.6% and 26.5%, respectively [23]. The increasing number of refugees, coupled with inadequate resources, raises critical questions about the capacity of the existing health system to effectively diagnose, manage, and treat these NTDs. Moreover, comprehensive data on NTDs and their prevalence are scarce in regions such as southwestern Uganda. Uganda’s national strategy for NTDs involves integrated control programmes [24]. This study assessed the capacity of primary healthcare centers to timely diagnose, treat, and manage soil-transmitted helminths and schistosomiasis among refugees in Nakivale Refugee Settlement, Southwestern Uganda, by looking at the availability of essential diagnostic and reporting infrastructure.

## Methods

### Study Design and Setting

This study employed a quantitative, cross-sectional design focusing on assessing health facility capacity. The study was conducted in Nakivale Refugee Settlement, located in southwestern Uganda, a large settlement hosting a diverse refugee population.

### Study Population and Sampling

The study population comprised 8 health facilities within Nakivale Refugee Settlement. These facilities included 6 Health Centre III (HCIII) and 2 Health Centre IV (HCIV) facilities, representing key points of primary healthcare service delivery in the settlement. These facilities were purposely sampled, as they served as the first point of contact for patients within the settlement.

### Data Collection

Data were collected through a structured quantitative assessment form administered at each health facility over one month to evaluate capacity for NTD management. The form captured facility characteristics (type, location, and bed capacity), laboratory capacity (availability of a dedicated lab, technician, and microscope), and diagnostic resources (tests for intestinal schistosomiasis and STH, alongside clinical guidelines and protocols). Reporting infrastructure was assessed through the use of the Health Management Information System and the availability of reporting forms. The survey also documented essential medicines (praziquantel, albendazole, mebendazole, and ivermectin) and laboratory supplies required for diagnosis, including slides, centrifuges, filters, syringes, staining reagents, Kato-Katz templates, and related consumables.

### Data Analysis

Completed questionnaires were first logged into Microsoft Excel 2016 for data entry, cleaning, and coding to ensure accuracy and consistency, after which the dataset was exported to STATA 17.0 for analysis. Facility characteristics such as facility type and distance from the town council were analyzed as categorical variables and summarized using frequencies and percentages, while the number of hospital beds was treated as a continuous variable and summarized using means and ranges. Laboratory capacity (presence of a laboratory, technician, and microscope), diagnostic resources (availability of tests and diagnostic guidelines), reporting infrastructure (HMIS use and reporting forms), essential medicines, and laboratory supplies were all analyzed as binary categorical variables (present = 1, absent = 0) and summarized descriptively using frequencies and proportions. Multi-select categories, such as diagnostic tests and laboratory consumables, were further disaggregated to present the availability of each item across the eight facilities, allowing a detailed profile of service readiness.

### Ethics Statement

This study was conducted in accordance with the ethical principles outlined in the Declaration of Helsinki. Ethics approval was obtained from the Lacor Hospital Institutional Research Ethics Committee (Reference Number: LACOR-2024-362). Written informed consent was obtained from all participants prior to data collection. Participants were informed about the study objectives, procedures, potential risks and benefits, their right to decline or withdraw at any time without any consequence, and confidentiality measures were assured.

## Results

### Types and Distribution of Health Facilities

Table 1 summarizes the assessment of 8 health facilities within Nakivale Refugee Settlement. Of these, 6 were identified as Health Centre III (HCIII) facilities, and 2 were Health Centre IV (HCIV) facilities. Regarding geographical accessibility, 7 out of the 8 facilities were located more than 10 km from the town council, with only one Health Centre IV situated within 10 km. The average number of hospital beds varied by facility type. Health Centre III facilities reported an average of 23 hospital beds, while Health Centre IV facilities had a higher average of 50 hospital beds.

**Table 1:** Baseline Characteristics of Assessed Health Facilities (N=8)

| Characteristic | Category | Count | Percentage (%) |
| --- | --- | --- | --- |
| **Total facilities assessed** |  | 8 | N/A |
| **Facility Type** | Health Center III | 6 | 75 |
|  | Health Center IV | 2 | 25 |
| **Geographical Accessibility** | >10km from town council | 7 | 87.5 |
|  | Within 10km from town council | 1 | 12.5 |
| **Average Hospital Bed Capacity** | Health Center III | 23 beds | N/A |
|  | Health Center IV | 50 beds | N/A |
| **Laboratory Presence** | Dedicated Laboratory | 8 | 100 |
|  | Designated Lab Technician | 8 | 100 |
|  | Microscope availability | 8 | 100 |

All 8 assessed health facilities reported the presence of a dedicated laboratory and a designated lab technician for the diagnosis of Neglected Tropical Diseases. Furthermore, all 8 facilities confirmed the availability of a microscope indicating a foundational diagnostic capability.

### Diagnostic Tests and Guiding Tools

Table 2 shows characteristics regarding essential medicines for the management of STH and schistosomiasis, with the majority of facilities being well-stocked. Seven out of the 8 facilities reported having both Albendazole and Mebendazole available. One facility reported having only Albendazole. All facilities, however, lacked praziquantel and ivermectin.

**Table 2:** Critical Gaps in Essential NTD Diagnostics and Treatment Medications.

| Characteristic | Item | Facilities with Item | Percentage(%) |
| --- | --- | --- | --- |
| **Availability of Essential Medicines** | Albendazole/Mebendazole(both) | 7 | 87.5 |
|  | Albendazole only | 1 | 12.5 |
|  | Praziquantel | 0 | 0 |
|  | Ivermectin | 0 | 0 |
| **Availability of Key Lab supplies** | Centrifuge tubes | 8 | 100 |
|  | Syringes | 8 | 100 |
|  | Cellophane | 0 | N/A |
|  | Kato Katz templates | 0 | N/A |
|  | Urine dipsticks | 7 | 87.5 |
|  | Dyes | 5 | 62.5 |
|  | Spatula | 3 | 37.5 |
|  | Membrane filters | 2 | 25 |
|  | Iodine | 4 | 50 |
|  | Malachite Green | 1 | 12.5 |
|  | Glycerine solution | 1 | 12.5 |
| **Available Diagnostic Tests for Intestinal Schistosomiasis and STH** | Clinical Guidelines | 7 | 87.5 |
|  | Laboratory protocols | 6 | 75 |
|  | Bench aids | 5 | 62.5 |

The availability of specific diagnostic tests for intestinal schistosomiasis and STH showed some variation across facilities, but commonly included microscopes, dyes, and assay strips. In terms of tools to guide diagnosis, all facilities reported the presence of at least some of the listed items, such as clinical guidelines, laboratory protocols, and bench aids, though the specific combination of these tools differed between facilities.

### Reporting Systems

The assessment of reporting infrastructure revealed a notable gap. While 5 of the 8 facilities had reporting forms available for the Health Management Information System, 3 facilities reported that these forms were not available. This indicates an inconsistency in the capacity for systematic data collection and reporting on NTD diagnosis and treatment (Table 3).

**Table 3:** Inconsistent NTD Reporting Infrastructure and Geographical Accessibility Challenges.

| Characteristic | Category | Facilities | Percentage(%) |
| --- | --- | --- | --- |
| Availability of Reporting Forms | Reporting forms for HMIS are available | 5 | 62.5 |
|  | Reporting forms for HMIS not available | 3 | 37.5 |

## Discussion

This study identified four major gaps in the readiness of health facilities in Nakivale Refugee Settlement to provide comprehensive NTD services, which included inadequate diagnostic supplies despite the presence of basic laboratory infrastructure, absence of essential medicines such as praziquantel and ivermectin, weak reporting systems, and geographical inaccessibility of most facilities. These limitations constrain the effectiveness of NTD prevention, diagnosis, and management, despite some encouraging strengths such as universal availability of laboratories, technicians, and microscopes.

Whereas all the facilities assessed reported having a designated laboratory technician and a functional microscope, critical diagnostic supplies were either inconsistently available or completely absent. When compared to other studies done in similar remote settings, the stronger baseline diagnostic infrastructure was an improvement, unlike similar studies in Tanzania and Burundi, where laboratory capacity was far more limited (44-45). However, although centrifuge tubes and syringes were available in all facilities, essential tools for WHO-recommended procedures such as urine sedimentation, centrifugation, and the Kato–Katz method (e.g., Kato-Katz templates, cellophane, and malachite green) were unavailable. This pattern mirrors findings from Tanzania, where facilities relied heavily on urine dipsticks and direct smear microscopy due to similar supply gaps (44). While urine dipsticks and direct smears are useful for rapid screening, their low sensitivity undermines accurate disease burden estimation, especially in high-risk refugee populations.

There was a noted absence of praziquantel and ivermectin across all facilities, despite their inclusion in the WHO Model List of Essential Medicines for schistosomiasis and onchocerciasis control (45). In contrast, albendazole and mebendazole were widely available (87.5%), reflecting an imbalance in NTD drug procurement. This finding is particularly concerning given the high prevalence of schistosomiasis in refugee-hosting regions of Uganda (46). Similar stock-outs have been documented in other low-resource settings, where fragmented supply chains and donor-driven drug distribution lead to inconsistent medicine availability (47). Without praziquantel and ivermectin, facilities in Nakivale Refugee Settlement remain unable to deliver integrated, guideline-based NTD care.

There were inconsistencies with the reporting infrastructure, with only 62.5% of facilities having HMIS reporting forms. This inconsistency limits systematic data capture, surveillance, and accountability. Similar reporting gaps have been observed in other sub-Saharan African health systems, where underreporting of NTD cases undermines programmatic planning and resource allocation (48). Strengthening reporting mechanisms is particularly critical in refugee settlements, where disease dynamics differ from national averages due to overcrowding, poor sanitation, and frequent mobility.

With regards to facility location, 87.5% of facilities were located more than 10 km from the town council, with only one HCIV within 10 km. This distance barrier adds to the structural challenges refugees face in accessing care. Previous studies have consistently highlighted that increased distance to health facilities reduces care-seeking behaviour, increases delays, and contributes to poor adherence to long-term treatment regimens (48). For NTDs that require periodic preventive chemotherapy, such barriers undermine programme reach and effectiveness.

This study’s strengths include its ability to identify the available diagnostic infrastructure in a refugee setting; however, the use of a single refugee settlement as a study site limits generalizability to other refugee settings. Availability of supplies and equipment was based on staff reports rather than direct observation or inventory checks, introducing a risk of reporting bias. The study also did not assess the quality of diagnostic practices or adherence to guidelines in daily use.

Future qualitative studies could explore the reasons behind the lack of specific laboratory consumables and reporting forms, as well as the challenges faced by health facility staff in maintaining these resources. Investigating the impact of facility distance on patient access to NTD services would also provide valuable insights.

In conclusion, this assessment shows that while Nakivale Refugee Settlement’s health facilities have a solid foundation for NTD management with labs, microscopes, and essential medicines in place, critical gaps remain. The lack of specific diagnostic supplies like Kato-Katz templates and cellophane, gaps in reporting systems, and the remote location of most facilities all pose significant challenges to effective NTD control. To address these, a regular supply of key laboratory consumables should be ensured through quarterly procurement reviews and better stock management. Strengthening and digitizing NTD reporting systems is also essential to improve surveillance and planning. Practical solutions to overcome remoteness, such as mobile outreach clinics or community-based diagnosis, should be prioritized.

Finally, future research should include qualitative studies to explore how distance and other barriers affect service use. Such insights will help tailor interventions to strengthen service delivery and accelerate progress toward NTD elimination in this vulnerable setting.

## List of Abbreviations

NTDs: Neglected Tropical Diseases
PHC: Primary Healthcare
WHO: World Health Organization
HCIII: Health Centre III
HCIV: Health Centre IV
STH: Soil-Transmitted Helminths
SDG: Sustainable Development Goal
USAID: United States Agency for International Development

## Declarations

### Ethics approval and consent to participate

Following the Declaration of Helsinki, Ethics approval for this study was obtained from the Lacor Hospital Institutional Research Ethics Committee, Reference Number [LACOR-2024-362]. Informed consent was obtained from all participants before data collection.

### Clinical trial number

Not Applicable

### Consent for publication

Not Applicable

### Availability of data and materials

The datasets generated and/or analyzed during the current study are not publicly available due to the sensitive nature of participant data and to maintain participant anonymity, but are available from the corresponding author on reasonable request.

### Authors’ contributions

TE conceived the study, participated in its design and coordination, and helped to draft the manuscript. AP and MPD participated in the study’s design, performed the statistical analysis, and drafted the manuscript. NMS, AC, MC, SK, AP, and MPD participated in data collection and interpretation. All authors read and approved the final manuscript

## Acknowledgements

The authors would like to thank the health workers in Nakivale Refugee Settlement for their participation in this study. We also extend our gratitude to Medical Teams International for their assistance during data collection. We also extend gratitude to the RSTMH Early Career Grant funding that facilitated the research process

---

**Figure 2:**
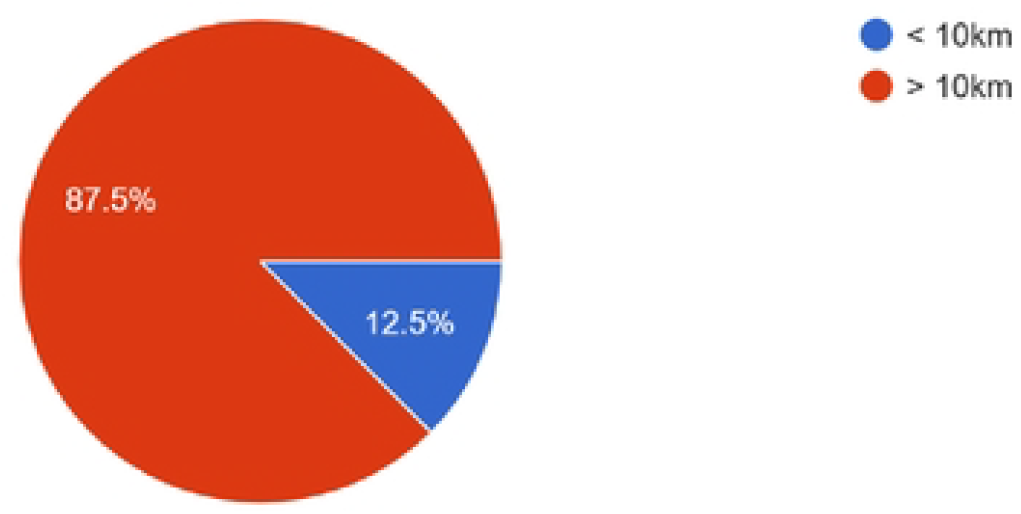
Distance of Health Facilities from Town Council.

**Figure 3:**
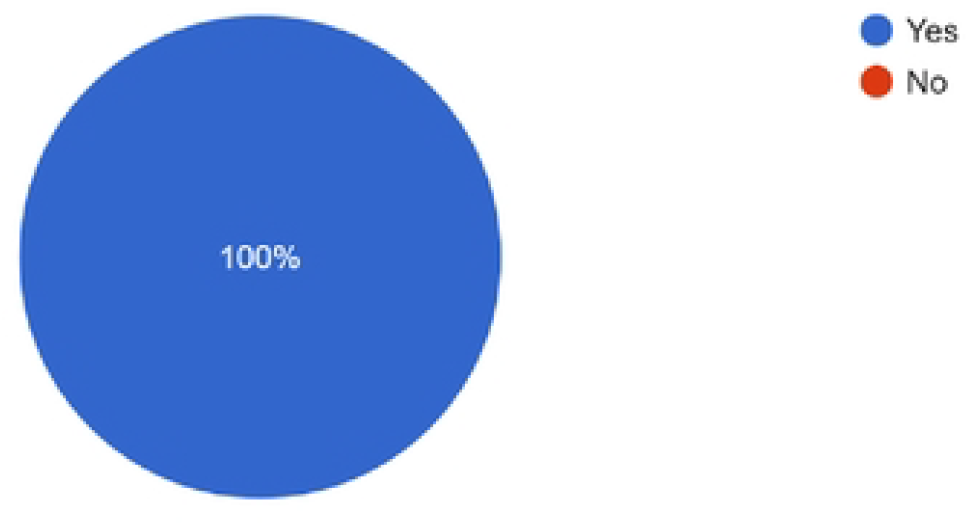
Availability of Microscopes.

**Figure 4:**
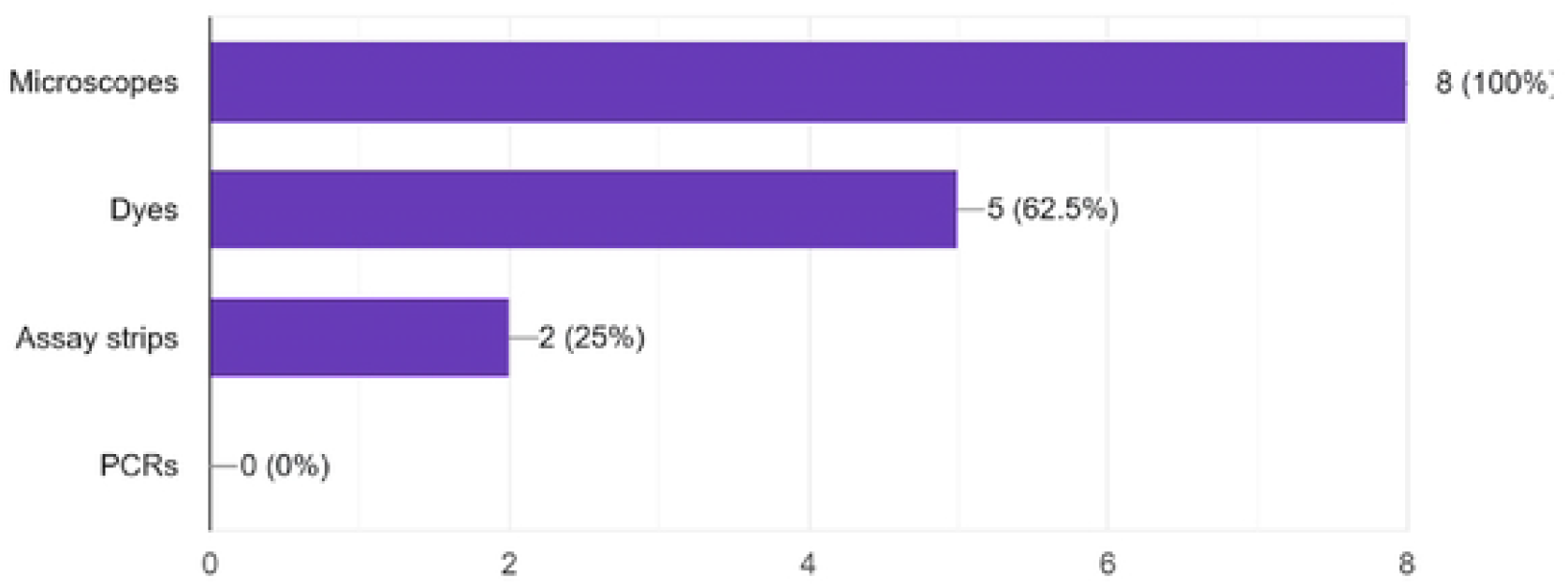
Available Diagnostic Tests for Intestinal Schistosomiasis and STH.

**Figure 5:**
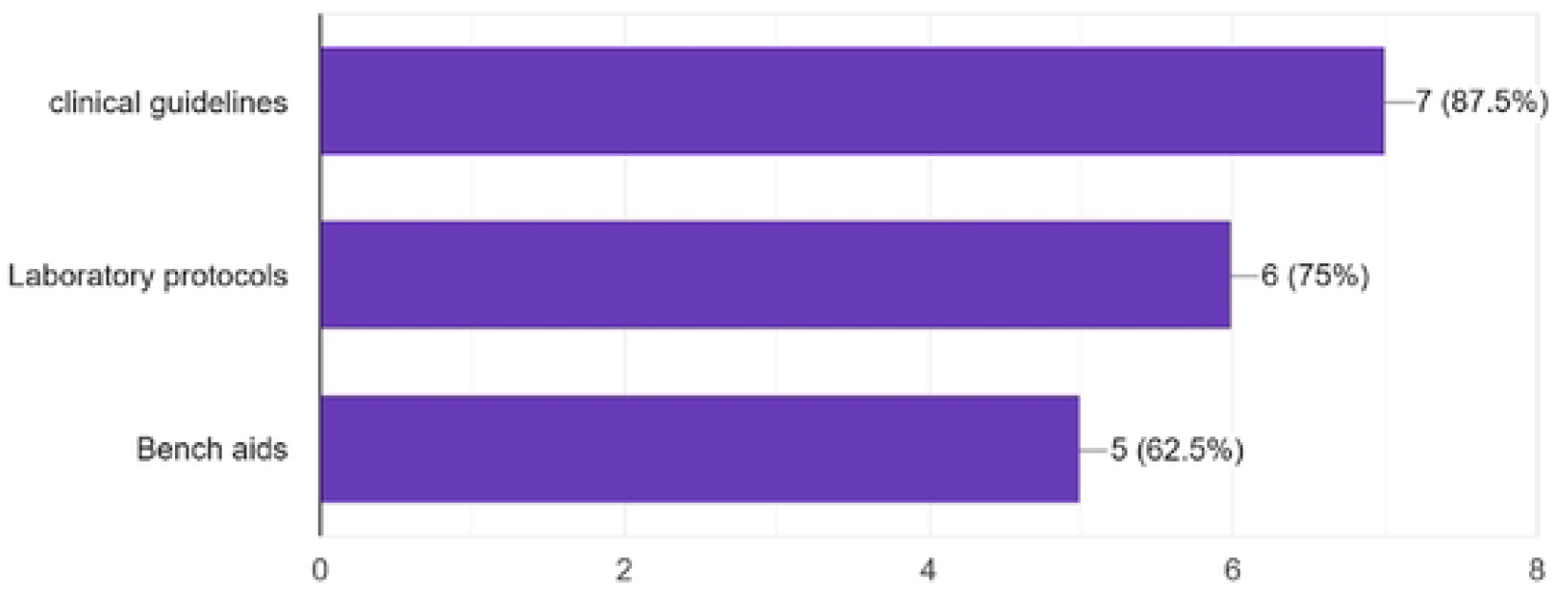
Available Diagnostic Tools for NTDs.

**Figure 6:**
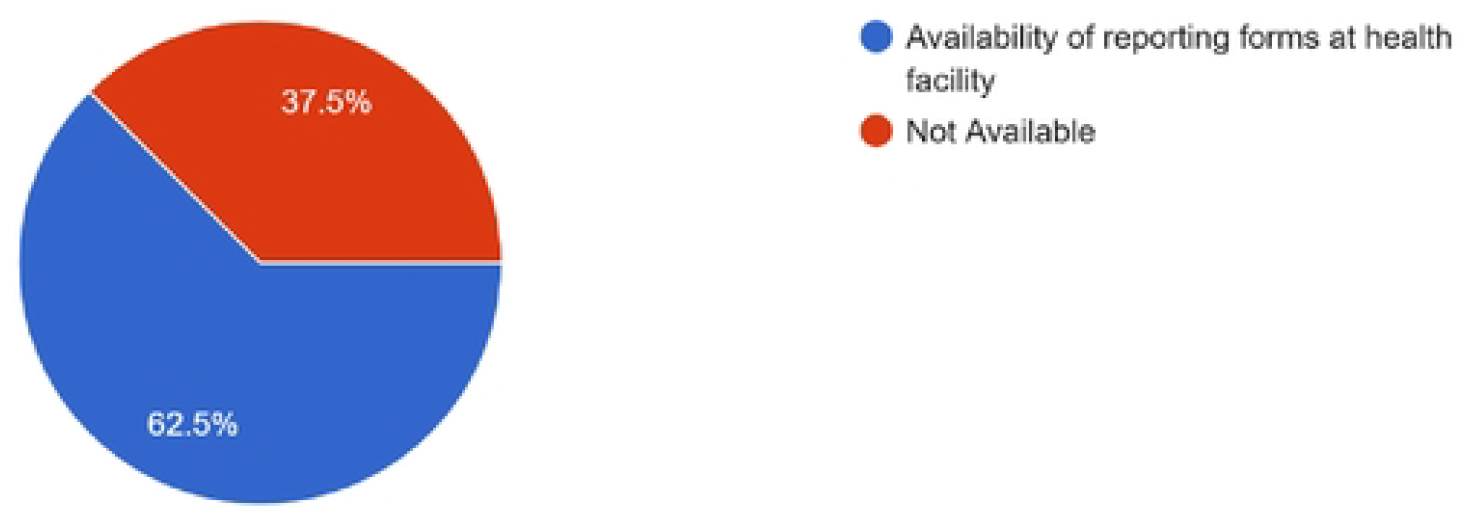
Availability of Reporting Forms.

**Figure 7:**
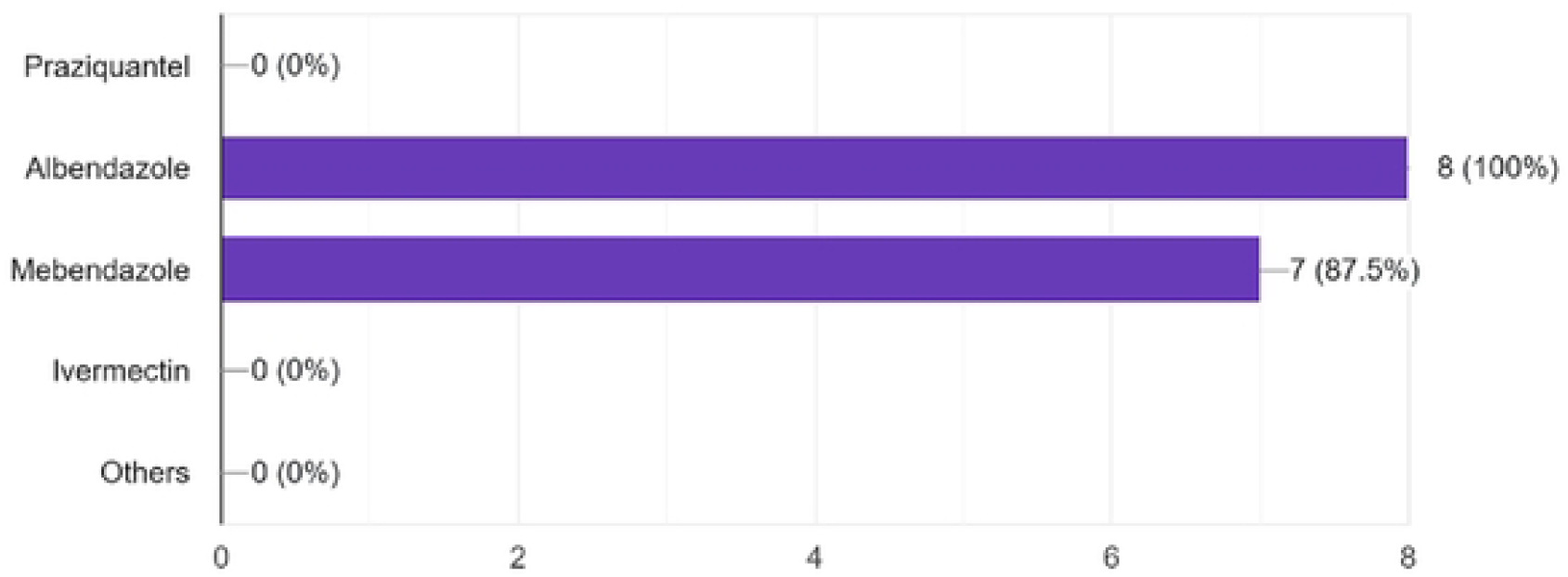
Availability of Essential Medicines.

**Figure 8:**
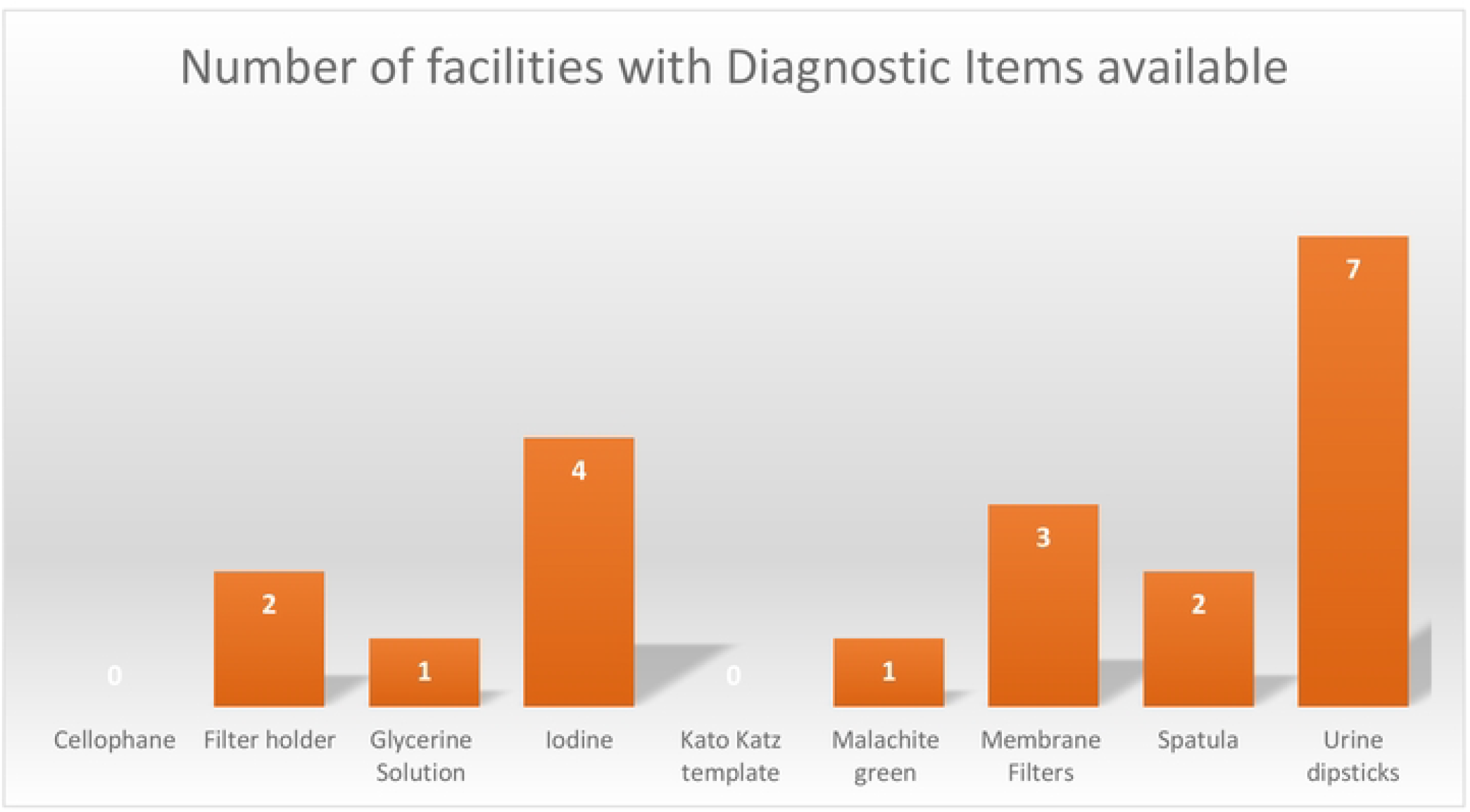
Availability of Laboratary Equipment.

